# Baseline Assessment of Drug-Drug Interaction Knowledge Among Healthcare Providers in Kibaha, Tanzania

**DOI:** 10.64898/2026.04.11.26350082

**Authors:** Ally Salim, Megan Allen, Kelvin Mariki, Thomas Pallangyo, Rose Lawrence Maina, Fatma Ally, Moses Minja, Kelvin Msovela, Jafary Liana

## Abstract

In the context of global health, the ability of frontline primary health providers to identify potential **Drug-Drug Interactions (DDIs)** is a critical component of patient safety. This is particularly true in settings like Tanzania, where drug dispensers often serve as the primary point of contact for patients. In this study, we establish a baseline for drug decision-making capabilities across multiple cadres of healthcare providers in Kibaha, Tanzania. We specifically distinguish between the ability to recognize **safe** drug combinations versus **harmful** ones. The findings reveal a critical asymmetry in provider performance: while professional training improves the recognition of safe combinations, it provides no advantage over lay intuition (and in some cases, a significant disadvantage) in detecting potentially harmful interactions.

## 1. Introduction

### 1.1 Healthcare Landscape

Like many countries globally, Tanzania faces significant health challenges driven by a shortage of healthcare workers. The physician-to-patient ratio is approximately 1:20,000, well below the World Health Organization recommended standard of 1:10,000 (Buguzi, 2017). Furthermore, three-fourths of doctors practice in urban areas, leaving rural populations with limited access to specialized care (Ministry of Health and Social Welfare, 2014). In this context, drug dispensers, Community Health Workers, and other front-line health providers play a crucial role as the first points of care for a majority of the population. They provide (differential) diagnoses, treatment recommendations, and medication dispensing. Despite their critical role, training levels vary significantly; while some possess formal qualifications, others rely on apprenticeship-based knowledge or learning on the job.

### 1.2 Clinical Decision Making and the Digital Gap

Decision-making among frontline providers in Tanzania relies predominantly on human memory and static paper-based guidelines. While the Ministry of Health provides Standard Treatment Guidelines and the National Essential Medicines List, adherence to these resources varies. In practice, dispensers often depend on recalled knowledge from prior training or interactions, which may be insufficient for complex cases involving multiple medications.

This stands in contrast to high-income countries, where Clinical Decision Support Systems (CDSS) are often deeply integrated into electronic health records and pharmacy dispensing software. In these settings, automated alerts for DDIs, contraindications, and dosage errors are standard safeguards that operate in real-time (Kuperman et al., 2007). In Tanzania, however, such digital safety nets are uncommon at the primary care level. While many have adopted electronic systems, they are often focused on administrative tasks rather than clinical decision support, leaving frontline dispensers to manage the cognitive load of prescribing (Mwogosi, 2025).

### 1.3 The Growing Challenge of Drug-Drug Interactions

Medication errors, including the failure to identify potential DDIs, are a major concern. As polypharmacy (the simultaneous use of multiple medicines) becomes more common, the cognitive burden on providers to memorize potential drug interactions increases. Recent data highlights the severity of this issue in the Tanzanian context. A study in rural Tanzania found that 33% of adult patients living with HIV were exposed to clinically relevant drug-drug interactions, with significant risks involving cardiovascular and analgesic co-medications (Schlaeppi et al., 2020). Pediatric patients are also affected, with dosing errors occurring in approximately 1 out of every 34 prescriptions at HIV clinics (Naik et al., 2017).

Overall, the recognition and appropriate management of DDIs is clearly suboptimal. This highlights a need for continuing education programs, ongoing pharmacovigilance activities, and the potential integration of effective digital decision support systems as treatment regimens become more complex

### 1.4 Study Aim

This study aimed to evaluate the baseline accuracy of drug dispensing decisions made by individuals ranging from laypersons to trained medical doctors. Specifically, we sought to:

- **Assess Baseline Accuracy:** Determine the current ability of providers to identify drug-drug interactions, distinguishing between “safe” and “unsafe” scenarios.
- **Compare Expertise:** Analyze performance differences across groups (Laypersons, Accredited Drug Dispensing Outlet (ADDO) Dispensers, Clinical Officers, Pharmacists, and Medical Doctors).
- **Inform Future Interventions:** Quantify the gap in medication safety to inform the development of future decision-support tools.

## 2. Study Objectives

The primary objective of this study was to measure the accuracy of DDI identification in a simulation-based environment. Secondary objectives included: 1) comparing decision-making accuracy across different levels of medical expertise, and 2) identifying specific gaps in knowledge regarding common drug combinations.

## 3. Methodology

### 3.1 Study Population

Data collection occurred in Kibaha, Tanzania from March 10-12, 2025. The study included 80 participants across five distinct groups: four professional groups and one layperson group.

Inclusion criteria required participants to be adults (18-45 years) with basic digital literacy. The groups and their characteristics were as follows:

- **Laypersons (n=30):** No medical training. Recruited from Kibaha Community Development College.
- **ADDO Dispensers (n=28):** Licensed/certified with >6 months dispensing experience.
- **Pharmacists (n=12):** Licensed with >6 months experience (combined Pharmacists and Techs).
- **Clinical Officers (n=5):** Licensed clinicians.
- **Medical Doctors (n=5):** Licensed with >1 year clinical experience.

### 3.2 Data Collection Process: The “Drug Cluster” Protocol

To simulate a dispensing environment without digital aids, we utilized a “Drug Cluster” station approach, where 2-3 medications were grouped into clusters, as seen in Figure 1.

**Figure 1:**
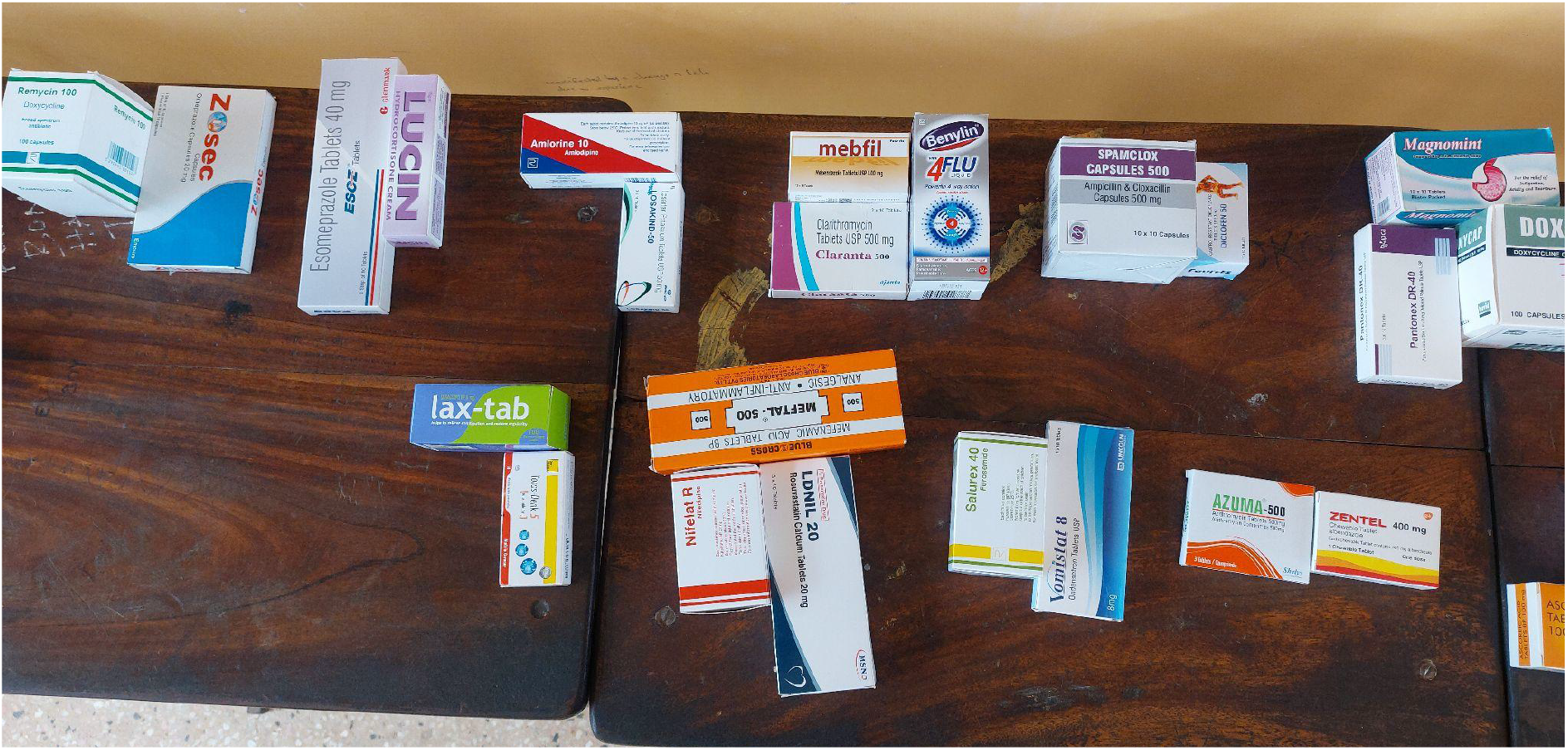
Clusters of drugs for participants to examine. Each cluster was numbered to act as a reference for the participant labeling.

Each cluster was a representation of a potential prescription or set of drugs that could be prescribed/ given to a patient. In total, there were 30 clusters to evaluate: 15 involved unsafe drug combinations that required the identification of potentially harmful interactions and 15 involved safe drug combinations requiring recognition of non-harmful pairings.

Participants were asked to review each cluster and determine whether it was “safe” or “unsafe” to provide to a patient. They were allowed to physically handle the medication packages to investigate ingredients. Medication information packets typically found inside the boxes were removed. After assessing a cluster, participants used a study data collection sheet to check the appropriate box indicating whether each of the clusters of medications were safe or not safe. Participants were prohibited from collaborating during the study.

Each cluster was designed before data collection by pharmacists optimizing for common medications the participants were likely to have encountered. Drug clusters with interactions were categorized as either major or moderate interactions.

### 3.3 Statistical Analysis

We used Bayesian inference to compare drug-drug interaction identification accuracy between healthcare provider groups and laypersons. We analyzed the “safe” and “unsafe” question types separately to distinguish between the cognitive tasks of danger detection and safety recognition.

For each group, we modeled the number of correct responses using a Binomial likelihood with *uninformative Beta(1,1)* priors on the probability of correct response. This allowed the data to drive posterior estimates without imposing prior assumptions about group performance. We computed posterior distributions for each group’s accuracy and for the difference between each professional group and laypeople.

To quantify evidence for or against group differences, we calculated Bayes factors using the Savage-Dickey density ratio (Faulkenberry, 2020), with the heuristic BF_10_ > 3 indicating substantial evidence for a difference and BF_01_ > 3 indicates substantial evidence for equivalence (Stefan et al., 2019). We report 95% highest density intervals (HDIs) for all parameters. To assess practical significance, we conducted Region of Practical Equivalence (ROPE) analysis with bounds of ±0.05, treating differences smaller than 5 percentage points as practically negligible. All models were implemented in PyMC (version 5.x) (PyMC Project Website) with 4 chains of 3000 samples each.

## 4. Results

The analysis revealed distinct patterns of performance depending on whether the scenario involved a safe or unsafe combination of drugs. Figure 2 shows the estimated probability of a correct response for each provider group, separated by interaction type. We see a clear divergence: <u>professionals significantly outperform laypersons in identifying safe combinations (green bars), but they fail to outperform the untrained laypersons in detecting unsafe ones (red bars).</u> The error bars represent the 95% Highest Density Intervals (HDI) for each estimate.

**Figure 2:**
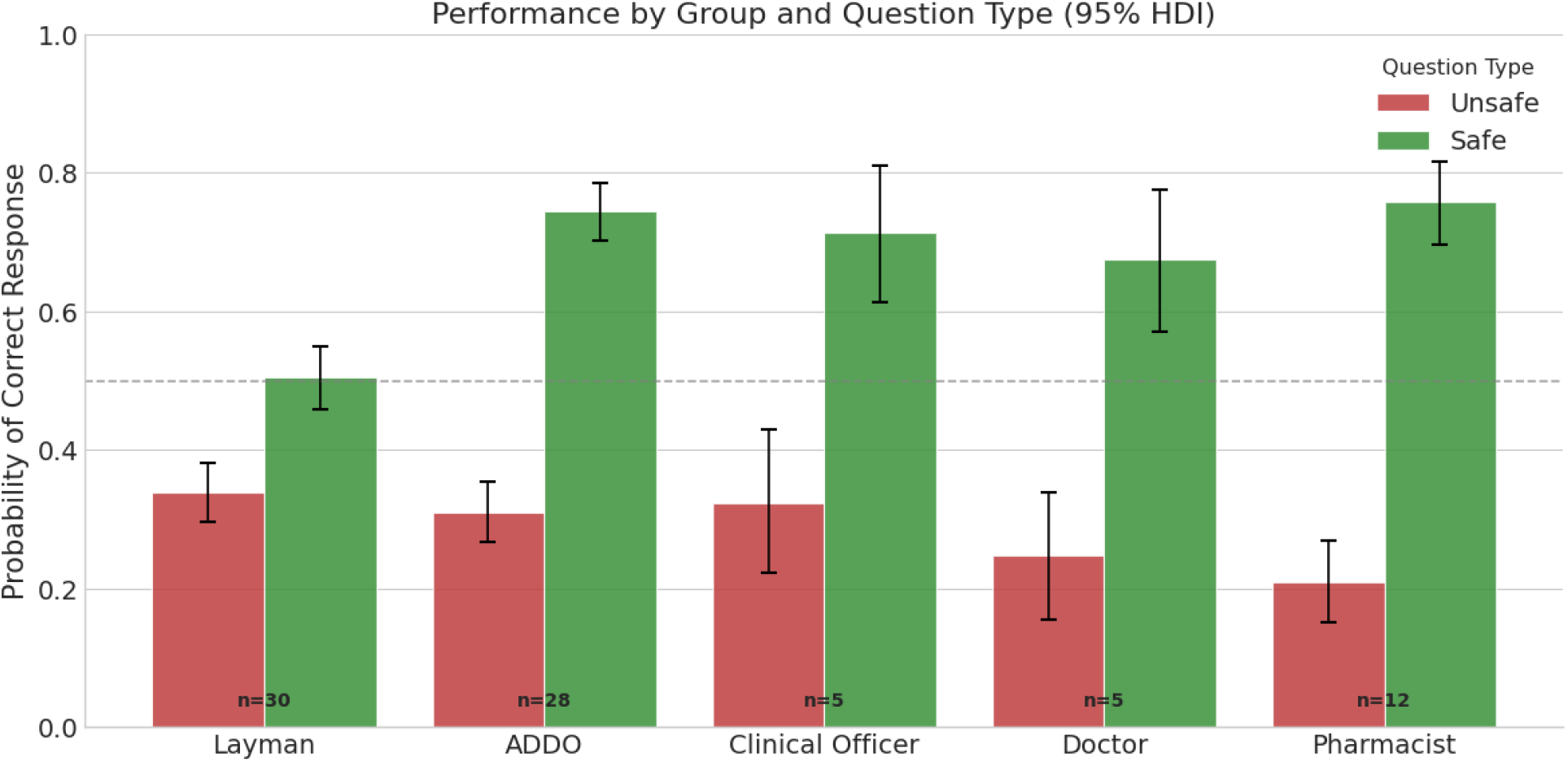
Probability of correct response by group, stratified by Safe vs. Unsafe interactions. Error bars represent 95% Highest Density Intervals (HDI).

### 4.1 Posterior Probability Estimates (Absolute Performance)

#### Unsafe Interactions (Danger Detection)

Performance was generally low across all groups, with mean accuracy ranging from 21% (pharmacists) to 34% (untrained laypersons). The density plots in Figure 4 (A) show high overlap among all groups, indicating that *<u>no specific group demonstrated a distinct advantage in identifying potential drug-drug interactions</u>*. Laypeople achieved a mean accuracy of 33.8% (95% HDI: 29.8%, 38.1%).

**Figure 4:**
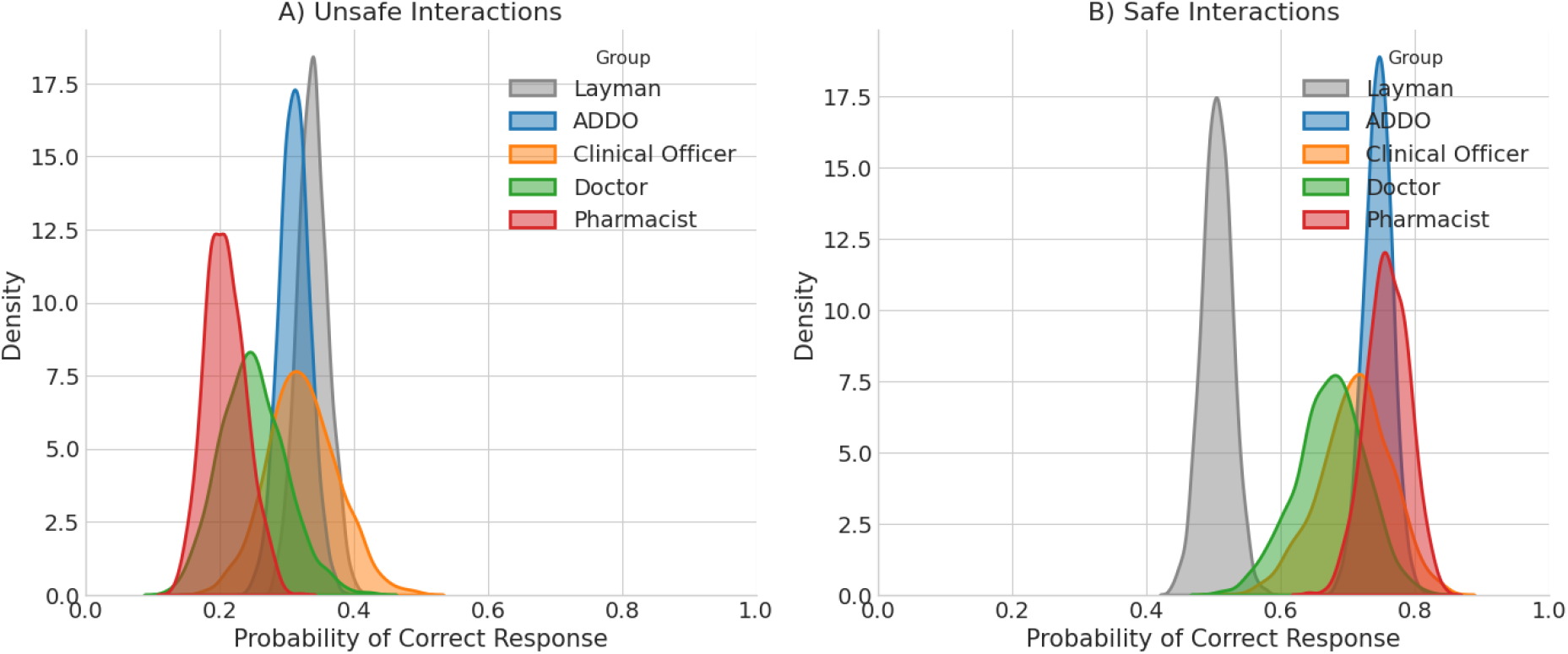
Posterior density plots of correct response probability. Panel A (left) shows the accuracy and high overlap among groups for Unsafe Interactions.

**Figure 5:**
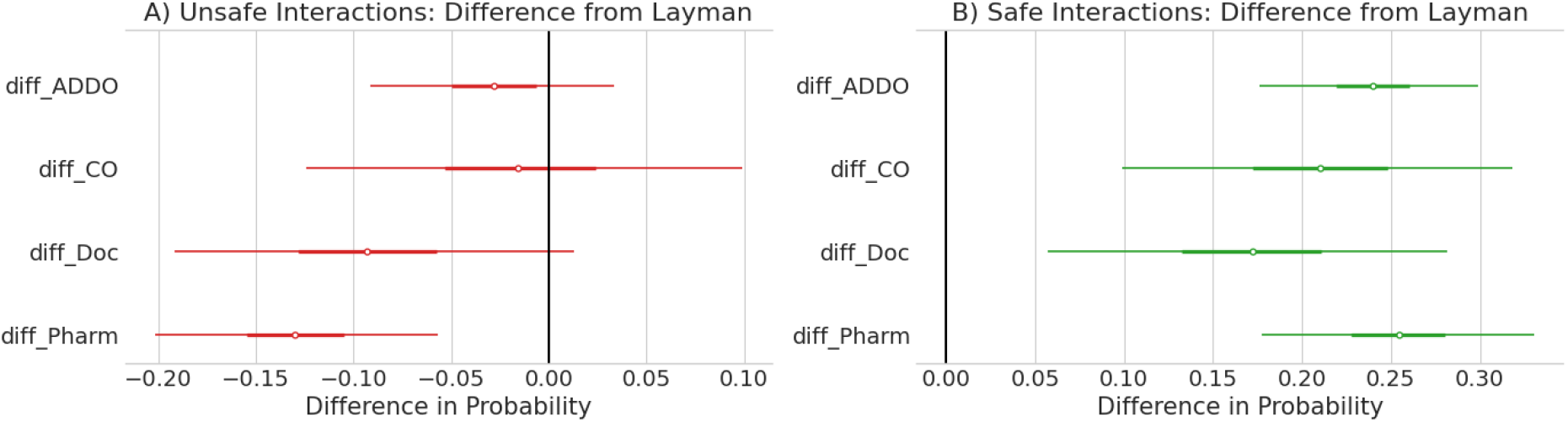
Forest plot showing the difference in probability of a correct response between professional groups and laypersons. Panel A (left) shows the relatively similar performance in recognizing unsafe interactions. Panel B (right) demonstrates the consistent superiority of professionals in recognizing safe interactions.

**Figure 6:**
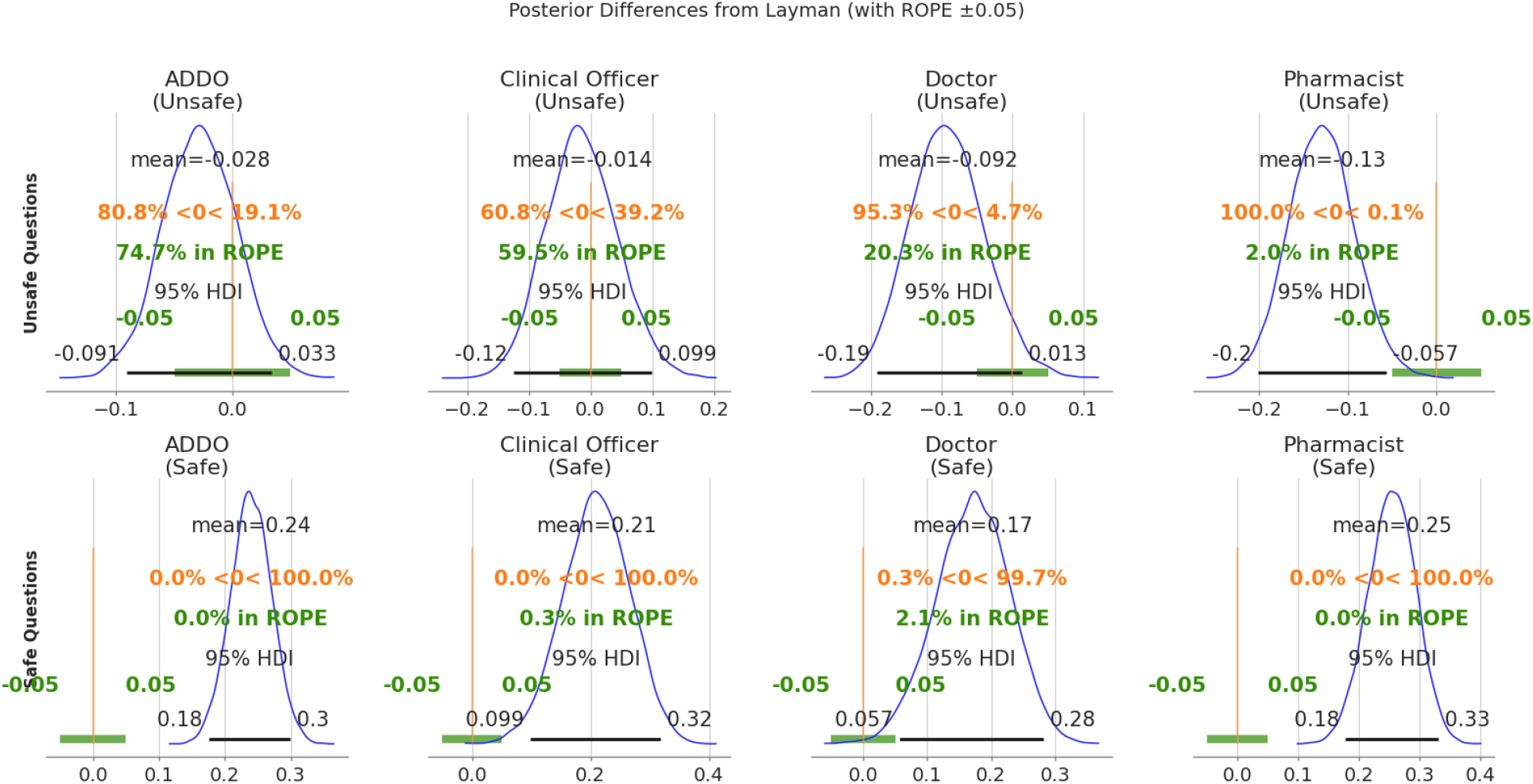
Detailed posterior differences relative to laypeople with Region of Practical Equivalence (ROPE) ±0.05. The top row illustrates that for Unsafe questions, professional performance falls largely within the ROPE (ADDO Dispenser) or significantly below it (Pharmacist).

#### Safe Interactions (Safety Recognition)

Figure 4 (B) reveals a clear separation in performance. While laypersons performed at chance level (Mean: 50.4%, 95% HDI: 45.9%, 54.6%), professional groups consistently achieved higher accuracy, ranging from 67% to 76%.

### 4.2 Comparative Performance Relative to Laypersons

#### Unsafe interactions

For the clusters with unsafe interactions, no professional group demonstrated superior ability to detect dangerous interactions compared to laypeople.

- **ADDO Dispensers**: Performed equivalently to laypeople (Mean difference: −0.028, 95% HDI: −0.091, 0.033), with substantial evidence for equivalence (BF_01_ = 8.72) and 74.7% of the posterior falling within the ROPE.
- **Pharmacists**: Demonstrated significantly lower accuracy than laypersons (Mean difference: −0.129, 95% HDI: −0.202, −0.057). The Bayes factor (BF_10_ = 16.1) indicates strong evidence for this negative difference, with 100% of the posterior distribution below zero.

#### Safe Interactions

In contrast, Panel B demonstrates that all professional groups <u>substantially outperformed laypeople</u> in recognizing safe combinations.

- **Pharmacists**: Showed the strongest performance over laypersons (Mean difference: +0.254, 95% HDI: 0.178, 0.330).
- **ADDO Dispensers**: Showed performance at par with pharmacists over laypersons (Mean difference: +0.240, 95% HDI: 0.176, 0.299).
- **Clinical Officers & Doctors**: Both groups performed significantly better than laypersons, with mean differences of +0.210 and +0.171, respectively.

All professional comparisons for safe questions shows clear evidence of superior performance (BF_10_ > 8), with >99% of the posterior distributions falling above zero and <3% in ROPE. These effects remained robust despite the limited sample sizes for the clinical officers(n=5) and the doctors(n=5).

## 5. Discussion

### 5.1 The Asymmetry of Expertise

Our findings contribute to the establishment of a baseline for identifying DDIs in the specific context and aims to provide prior probabilities for future work. We highlight the importance of separately evaluating decisions regarding potentially harmful versus safe drug interactions, as this enhances our understanding of how different healthcare providers perform relative to their expertise. Interestingly, we found that while professional training enhances recognition of safe drug combinations, it provides no advantage (and in the case of pharmacists, a disadvantage) in identifying harmful interactions.

Several mechanisms may explain this. First, pharmaceutical education may emphasize approved drug combinations and standard treatment protocols rather than comprehensive memorization of contraindicated pairings. Similarly, clinicians learn what they *can* prescribe together; dangerous combinations may be addressed through reference tools rather than recall. Second, response bias may contribute: laypersons may default to “unsafe” when uncertain, a cautious heuristic that inflates unsafe question scores while depressing safe question performance. Healthcare providers, confident in their training, may be more willing to clear combinations as safe, a pattern that serves them well for genuinely safe pairings but poorly when danger recognition is required.

It’s important to note that drug dispensers in this context rarely ask a patient or client what medication they are currently taking, and given our findings, that information might not be as useful as we’d hope.

### 5.2 Unexpected Pharmacist Paradox

The pharmacist finding was unexpected and requires more study and attention with larger sample sizes. Their poor performance on unsafe questions (21% vs. 34% for laypersons), combined with their strong performance on safe questions (76%), may suggest a pronounced overconfidence in drug safety. This pattern may reflect occupational context: pharmacists in community settings routinely approve medication combinations, potentially developing a bias toward clearance. The clinical implications are concerning, as pharmacists often serve as the final checkpoint before patients receive potentially interacting medications.

### 5.3 Limitations

Sample sizes for clinical officers (n=5) and doctors (n=5) were small, resulting in wide credible intervals and limited power to detect differences. While Bayesian methods appropriately quantify this uncertainty, replication with larger samples is needed. Additionally, the study was conducted in Kibaha, Tanzania, and findings may not generalize to settings with different training curricula or drug formularies. The assessment used a binary correct/incorrect format; clinical practice involves graded assessments of interaction severity that may reveal different patterns of expertise.

## 6 Conclusion

In many countries such as Tanzania, the status quo relies on providers detecting drug-drug interactions via memory alone. This study demonstrates that while training and experience improve a provider’s ability to confidently clear safe medications, it offers no protection against potentially harmful errors compared to an untrained layperson. The finding that pharmacists were statistically more likely to miss a dangerous interaction than a random layperson is a stark indicator that human cognition is ill-suited for the complex combinatorial task of DDI screening. Implementing digital decision support systems that automatically flag these interactions is not merely an enhancement for frontline healthcare providers, but a necessary safety net for even the most highly trained professionals.

## Data Availability

All data produced in the present study are available upon reasonable request to the authors.

## References

Buguzi, S. (2017). Half of young doctors to quit by 2025: report. The Citizen. https://www.thecitizen.co.tz/News/Half-of-young-doctors-to-quit-by-2025--report/1840340-3856488-811sq9z/index.html

Faulkenberry, T. J. (2020, August 26). Savage-Dickey density ratio for computing Bayes factors. The Book of Statistical Proofs. https://statproofbook.github.io/P/bf-sddr.html

Human Resources Development Directorate, Ministry of Health and Social Welfare. (2014). Human resource for health country profile 2013/2014. Dar es Salaam, United Republic of Tanzania. ISBN: 978-9987-737-05-5

Kuperman, G. J., Bobb, A., Payne, T. H., Avery, A. J., Gandhi, T. K., Burns, G., Classen, D. C., & Bates, D. W. (2007). Medication-related clinical decision support in computerized provider order entry systems: a review. Journal of the American Medical Informatics Association : JAMIA, 14(1), 29–40. 10.1197/jamia.M2170

Mwogosi, A. (2025). Electronic health record systems in reducing medication and diagnostic errors in Tanzanian health care. DOI: 10.1108/IDD-12-2024-0195

Naik, N. M., Mbwanji, R. N., Mgawe, M., Ongoa, R., Gesase, A. E., Schutze, G. E., Minde, M., & Mwita, L. F. (2017). Pharmaceutical Dosing Errors at a Pediatric HIV Clinic in Mwanza, Tanzania. The Pediatric infectious disease journal, 36(10), 973–975. 10.1097/INF.0000000000001639

PyMC Project Website. https://www.pymc.io/welcome.html

Rutta, E., Liana, J., Embrey, M., Johnson, K., Kimatta, S., Valimba, R., Lieber, R., Shekalaghe, E., & Sillo, H. (2015). Accrediting retail drug shops to strengthen Tanzania’s public health system: an ADDO case study. Journal of pharmaceutical policy and practice, 8, 23. 10.1186/s40545-015-0044-4

Schlaeppi, C., Vanobberghen, F., Sikalengo, G., Glass, T. R., Ndege, R. C., Foe, G., Kuemmerle, A., Paris, D. H., Battegay, M., Marzolini, C., Weisser, M., & Kilombero and Ulanga Antiretroviral Cohort (KIULARCO) study group (2020). Prevalence and management of drug-drug interactions with antiretroviral treatment in 2069 people living with HIV in rural Tanzania: a prospective cohort study. HIV medicine, 21(1), 53–63. 10.1111/hiv.12801

Stefan, A. M., Gronau, Q. F., Schönbrodt, F. D., & Wagenmakers, E.-J. (2019). A tutorial on Bayes Factor Design Analysis using an informed prior. Behavior Research Methods, 51(3), 1042–1058. 10.3758/s13428-018-01189-8

